# Strengthening health system’s capacity for pre-exposure Prophylaxis for adolescent girls and young women and adolescent boys and young men in South Africa (SHeS’Cap – PrEP): Protocol for a mixed methods study in KwaZulu-Natal, South Africa

**DOI:** 10.1101/2022.02.18.22271171

**Authors:** Edward Nicol, Trisha Ramraj, Mbuzeleni Hlongwa, Wisdom Basera, Ngcwalisa Jama, Carl Lombard, Tracy McClinton-Appollis, Darshini Govindasamy, Desiree Pass, Noluntu Funani, Sarah Aheron, Ariana Paredes-Vincent, Jennifer Drummond, Mireille Cheyip, Sibongile Dladla, Jason Bedford, Cathy Mathews

## Abstract

**Introduction:** Pre-exposure prophylaxis (PrEP) is an effective prevention intervention that can be used to control HIV incidence especially among people who are at increased risk for HIV such as adolescent girls and young women (AGYW) and adolescent boys and young men (ABYM). In South Africa, various approaches of delivering PrEP have been adopted at different service delivery points (facility-based only, school-based only, community-based only and hybrid school-facility and community-facility models) to overcome challenges associated with individual, structural, and health systems related barriers that may hinder access to and uptake of PrEP among these populations. However, little is known about how to optimize PrEP implementation and operational strategies to achieve high sustained uptake of good quality services for AGYW and ABYM. This study aims to identify effective and feasible PrEP models of care for improving PrEP uptake, continuation, and adherence among AGYW and ABYM.

**Methods and analysis:** A sequential explanatory mixed-methods study will be conducted in 22 service delivery points (SDPs) in uMgungundlovu district, KwaZulu-Natal, South Africa. We will recruit 600 HIV negative, sexually active, high risk, AGYW (aged 15-24 years) and ABYM (aged 15-35 years). Enrolled participants will be followed up at 1-, 4- and 7-months to determine continuation and adherence to PrEP. We will conduct two focus group discussions (with 8 participants in each group) across four groups (i. Initiated PrEP within 1 month, ii. Did not initiate PrEP within 1 month, iii. Continued PrEP at 4/7 months and iv. Did not continue PrEP at 4/7 months) and 48 in-depth interviews from each of the four groups (12 per group). Twelve key informant interviews with stakeholders working in HIV programs will also be conducted. Associations between demographic characteristics stratified by PrEP initiation and by various service-delivery models will be assessed using Chi-square/Fishers exact tests or t-test/Mann Whitney test. A general inductive approach will be used to analyze the qualitative data.

**Ethics and dissemination:** The protocol was approved by the South African Medical Research Council Health Research Ethics Committee (EC051-11/2020). This project was reviewed by the U.S. Centers for Disease Control and Prevention (Atlanta, GA), Centers for Global Health Associate Director for Science in accordance with CDC human research protection procedures and was determined to be research, but CDC investigators did not interact with human subjects or have access to identifiable data or specimens for research purposes. Provincial and district level approval has been granted. Findings from the study will be communicated to the study population and results will be presented to stakeholders and at appropriate local and international conferences. Outputs will also include a policy brief, peer-reviewed journal articles and research capacity building through research degrees.

## Introduction

South Africa had an estimated 7.7 million people living with HIV (PLHIV) in 2018, with HIV prevalence in the general population at 20.4% (1, 2). In 2018, there were 240,000 new HIV infections (1, 2), placing South Africa at an annual incidence of 4.94 per 1,000 uninfected adults (2). Young women aged 15-24 years accounted for 37% of new HIV infections in South Africa in 2017 (3, 4). In 2018, 33,000 adolescent girls were diagnosed with HIV compared to only 4,200 adolescent boys (1).

Several factors account for the increased vulnerability of adolescents and young women (AGYW) to HIV infection including intergenerational sexual relationships, gender-based violence, exclusion from economic opportunities, and a lack of access to secondary school (5, 6). Poverty and the gender power inequalities also increase the vulnerability of AGYW to HIV (7, 8). Relationships with older men may lead to power imbalances which increase the risk of intimate partner violence (IPV) victimisation for AGYW, and decrease the chance of knowing a partner’s HIV status or using condoms (6).

The World Health Organisation (WHO) recommended that people at substantial risk of HIV i.e., HIV incidence of 3 per 100 person-years or higher in the absence of pre-exposure prophylaxis (PrEP) – should also be offered PrEP within a comprehensive HIV package (9). South Africa aims to reduce the annual number of new infections in the general population to under 100,000 by 2022 (10). To achieve this, the country is currently implementing several HIV prevention strategies including PrEP. The South African National Department of Health (SANDoH) approved PrEP in 2015 with roll-out starting in 2016 (11).

The use of PrEP is potentially hindered by many factors including poor awareness, stigma, discrimination, and inadequate supplies and stock-outs at health facilities (12, 13). Among university students in South Africa, only one in five were aware of PrEP. While those who tested for HIV were significantly more likely to be aware of PrEP, only adequate family support and having discussed HIV/STIs with sexual partners were associated with increased awareness on PrEP (3). Adolescents and young adults are often reluctant to seek sexual and reproductive health services due to fear or experiences of poor service delivery, stigma, and discrimination from health care providers (13-16).

While AGYW are disproportionately affected by HIV, heterosexual men remain a critical population in HIV prevention. Men have lower HIV testing, linkage and retention to care rates compared to women (17). While 78% of men living with HIV know their HIV status, compared to 89% of women in South Africa, only 67% of men diagnosed with HIV are on antiretroviral therapy (ART), compared with 72% of women diagnosed with HIV (18). Lack of linkage and retention to ART contributes to increased virological failure which may contribute to increased mortality rates among men, further widening the life expectancy gap between men and women (19). Many barriers have been reported to deter men from accessing HIV treatment services, including concerns about confidentiality, long queues, inconvenient clinic operating hours, stigma and time and money spent travelling to seek care (20-24).

To address continued high HIV incidence among AGYW and adolescent boys and young men (ABYM), and the factors that increase their vulnerability to infection, various differentiated care models – tailored and streamlined client-catered HIV services – have been implemented throughout South Africa, including the Determined, Resilient, Empowered, AIDS-free, Mentored and Safe (DREAMS) initiative (25). These services comprise of adolescent and youth-friendly clinics (26), mobile clinics for reproductive health services for youth (26, 27) and community-based services (28), of which there is a high willingness among AGYW to initiate PrEP when delivered through an integrated approach within youth-friendly settings (26). AGYW who used contraception were significantly more likely to initiate PrEP on the same day compared to those who declined contraception (27).

While various approaches and innovations have been adopted at various service delivery points (SDPs), to improve the uptake of, and continuation on PrEP, there is lack of evidence exploring the different models of intervention designed to improve PrEP uptake and continuation. A comparative evaluation of service delivery models to determine which PrEP models of care are most successful in improving PrEP uptake and continuation is required, especially for at-risk populations such as AGYW and ABYM in South Africa. In this manuscript, we describe the SheS’Cap **–** PrEP protocol, a mixed methods study which aims to identify effective and feasible PrEP models of care for improving PrEP uptake, continuation, and adherence among AGYW and ABYM in the uMgungundlovu district, KwaZulu-Natal province, South Africa.

## Materials and methods

### The objectives of the study are

1. To assess the characteristics of AGYW (15-24 years) and ABYM (15-35 years) tested for HIV in the uMgungundlovu district who initiate PrEP compared to those who do not initiate PrEP and those who continue PrEP compared to those who discontinue PrEP.
2. To determine the rates of uptake of (initiation on) PrEP among AGYW and ABYM and continuation on PrEP at 1 (t_1_), 4 (t_3_) and 7 (t_6_) months.
3. To compare the rates of PrEP uptake and continuation of various existing PrEP-based HIV prevention service-delivery models. The service delivery models include facility-only, school-based only models, community-only and hybrid school-facility and community-facility models. The hybrid service delivery model is not an initial point of sample recruitment, as this is a model which we expect to emerge during the study based on client decision-making (e.g., an individual may initiate PREP at a facility, but continue care at a community service delivery point).
4. To investigate the social, economic, health system, and behavioural factors that facilitate or impede PrEP cascade outcomes among AGYW and ABYM in the uMgungundlovu district, including factors relating to the various models of PrEP service delivery and their acceptability.

### Setting

This study will be conducted in uMgungundlovu district in KwaZulu-Natal province, South Africa (Fig 1). We selected facilities in consultation with provincial and district Department of Health teams. uMgungundlovu is a high HIV prevalence district, accounting for 24% and 37% for people aged between 15-49 years among males and females, respectively (29). In KwaZulu-Natal, 70% of people living with HIV were on ART in 2017 (18).

**Figure 1:**
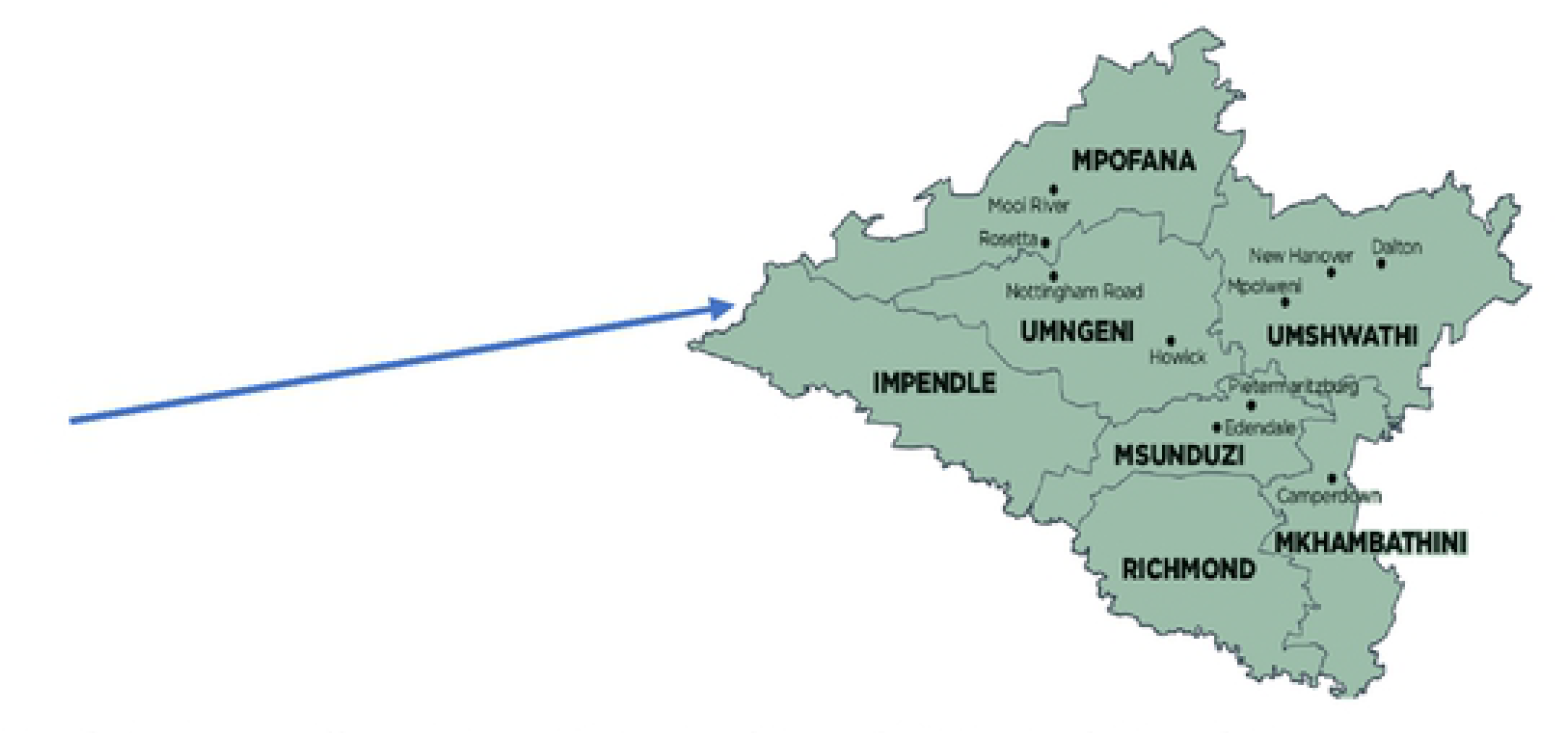
Sub-districts of uMgungundlovu District Municipality within Kwa-Zulu Natal Province *Source: File:Map of KwaZulu-Natal with municipalities named and districts shaded (2016). https://en.wikipedia.org*

The district is one of the National Health Insurance (NHI) pilot sites, which has 46 fixed clinics (primary health care facility or community health centre that has permanent staff and equipment and that provides an 8-24-hour service per day for five or more days a week), 17 mobile clinics (vehicles equipped with primary health care (PHC) provisions that transport health workers from a source facility to stopping points where they render their services) and one state aided clinic (private institution which is totally or partially funded by the State). The population residing in this province are predominantly poor and live in rural areas, and the majority (78%) use public health services (30).

### Study design

The SHeS’Cap-PrEP study is a sequential, explanatory, mixed-methods study that will be conducted at 22 service delivery points (SDPs). We will conduct a cross-sectional survey of AGYW and ABYM who tested HIV negative at the selected PrEP service delivery points and follow them up for up to 6 months after initiating PrEP, and a cohort study of those who initiate PrEP. We will also include a qualitative component, including in depth interviews (IDI) and focus group discussions to gather perspectives of participants from those who initiate and those who do not initiate PrEP at month 1 and from those who continue and those who do not continue PrEP at month 3 or 6. For the quantitative component, we will administer questionnaires and accessing routine service records at enrolment and at 1, 4, and 7 months to obtain HIV test results, and to describe PrEP initiation and continuation. Data collection will be over a 13-month period (July 2021 to July 2022). AGYW and ABYM will be recruited at selected PrEP SDPs (facility-, school- and community-based) and followed up for 7 months using quantitative and qualitative data collection methods.

### Population and recruitment

The target population for this study comprises sexually active AGYW aged 15 – 24 years and ABYM aged 15 – 35 years, who test HIV negative at 12 selected SDPs, in each of the seven sub-districts (strata) in the uMgungundlovu district. We will recruit participants who test HIV negative during the routine facility-based, school-based, and community-based HIV testing services. These cohorts will be recruited and monitored through surveys using questionnaires and routine pharmaceutical records of PrEP pill collection at one month, and SDP records at four and seven months. Under routine settings, patients are expected to be screened for HIV risk to qualify for PrEP. We will, however, not conduct screening for HIV risk to assess PrEP eligibility, as this is the responsibility of the HIV Testing Service (HST)/PrEP providing partner but will collect information on the participants’ risk using a PrEP risk assessment test at the baseline interview. Eligible participants will be considered based on the inclusion and exclusion criteria listed in Table 1.

**Table 1:**
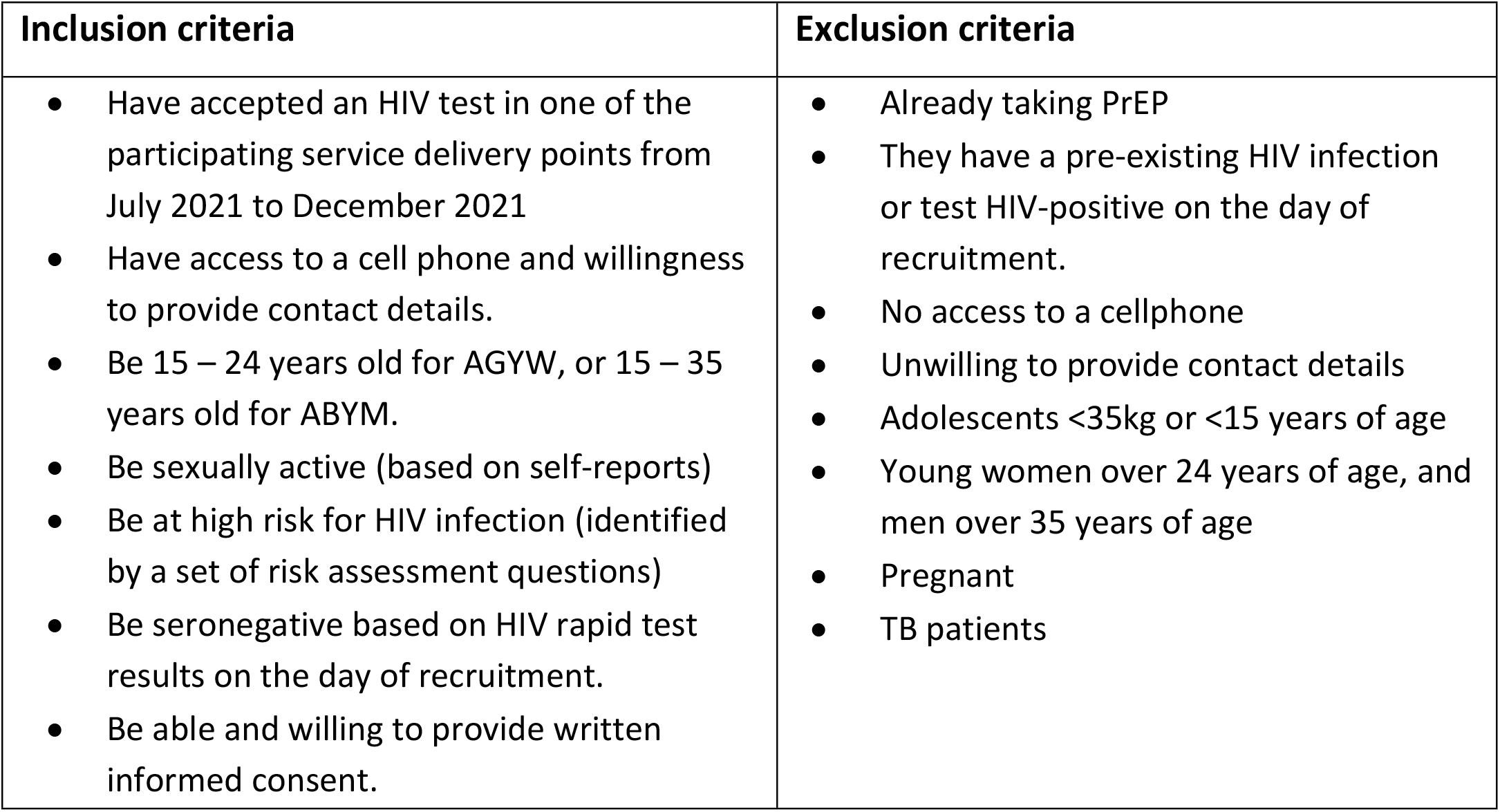
Eligibility criteria for the SHeS’Cap-PrEP study in uMgungundlovu district, KZN, 2021.

| Inclusion criteria | Exclusion criteria |
| --- | --- |
| • Have accepted an HIV test in one of the participating service delivery points from July 2021 to December 2021<br>• Have access to a cell phone and willingness to provide contact details.<br>• Be 15 – 24 years old for AGYW, or 15 – 35 years old for ABYM.<br>• Be sexually active (based on self-reports)<br>• Be at high risk for HIV infection (identified by a set of risk assessment questions)<br>• Be seronegative based on HIV rapid test results on the day of recruitment.<br>• Be able and willing to provide written informed consent. | • Already taking PrEP<br>• They have a pre-existing HIV infection or test HIV-positive on the day of recruitment.<br>• No access to a cellphone<br>• Unwilling to provide contact details<br>• Adolescents <35kg or <15 years of age<br>• Young women over 24 years of age, and men over 35 years of age<br>• Pregnant<br>• TB patients |

### Sampling

#### Selection of participating service delivery points

Excluding correctional facilities and non-medical sites, there are 64 public sector facilities providing PrEP in uMgungundlovu. These included 46 fixed clinics, 17 Mobile Clinics, and one state-aided clinic. To ensure feasibility of reaching enrolment, a threshold will be set to include only facilities which reported 10 or more patients testing positive per month (average over 12 months, September 2019-August 2020, DHIS data). The selection of the SDPs was based on the presence of an ongoing intervention (DREAMS core package of interventions in the region) amongst adolescent girls as well as the link between the primary health care facilities and schools/community college/men-linked services. Within each of the seven strata (sub-districts), 22 SDPs will be conveniently selected based on mean HIV positive rates in our target groups (AGYW/ ABYM) in each of the 64 facilities providing ART services in the district, and facilities that are linked with schools, community colleges and men-linked HIV services. Table 2 shows the different SDPs, highlighted in grey).

**Table 2:**
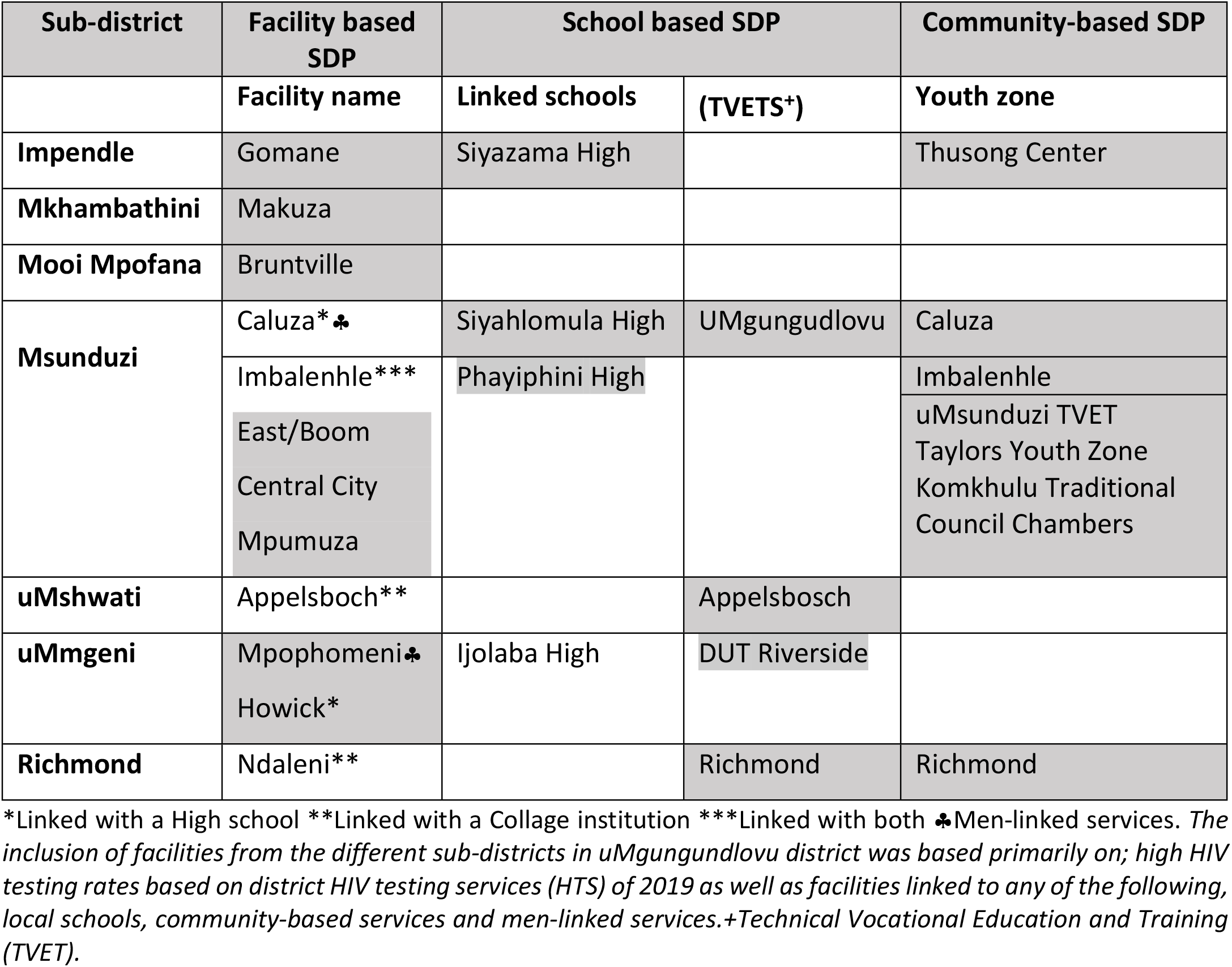
Service delivery points selected for inclusion in the SHeS’Cap-PrEP study in uMgungundlovu district, KZN, 2021.

### Sample size for individuals to be enrolled

Recent preliminary findings on uptake and adherence to PrEP in a sub-study nested within a clinical trial seeking to measure HIV incidence in Durban, South Africa estimated the rate of PrEP initiation to be 46% (31). We replicated the initiation rates of 46% in uMgungundlovu based on the similarity in populations. We assumed the initiation rates to be as low as 21% and to be as high as 46% based on the estimates from a similar population. Sample size calculations (Table 3) are illustrative of the power of the study design to detect difference in coverage by service delivery models and are based on the comparison of proportion between clusters of fixed size in each group at a specific time point (i.e., 6 months). The sample and power calculation were done using Stata v14.2 (32).

**Table 3:** Power and sample size for the comparison of service delivery model.

| Models | alpha | beta | k<sub>1</sub> | k<sub>2</sub> | m<sub>1</sub> | m<sub>2</sub> | delta | p<sub>1</sub> | p<sub>2</sub> | rho |
| --- | --- | --- | --- | --- | --- | --- | --- | --- | --- | --- |
| Community vs. Facility models | 0.05 | 0.80 | 4 | 4 | 50 | 50 | 0.25 | 0.21 | 0.46 | 0.04 |
| Community vs. School models | 0.05 | 0.80 | 4 | 4 | 20 | 20 | 0.25 | 0.21 | 0.46 | 0.02 |

Where *alpha* - type 1 error; *beta* - power; *m* – average cluster size; *k* - number of clusters; *delta* – difference between proportions; *p* – PrEP initiation estimate (proportion); *rho* – inter-cluster correlation.

For the quantitative component, we will enrol a sample size of 200 (50 participants x 4 SDPs) per service delivery model, fitting the proposed risk profile assessed at the baseline visit, to inform the overall number that may be reached during the seven months study period (600 plus additional 50 participants) to make provision for drop out and non-response. For the qualitative component, we will conduct interviews with 84 participants who initiated PrEP at 1 month in a service delivery point such as primary care clinic, school or community service and 28 participants who did not initiate PrEP. Based on the 1-month outcome data, we will purposively select 28 participants who successfully initiated PrEP, 28 who did not initiate PrEP, 28 participants who continued PrEP at four months and 28 who discontinued PrEP at four months. We will sample these participants from across the various service delivery models, based on age and gender. We will conduct two FGDs (with 8 participants in each group) and 12 IDIs from each of the four groups. We will conduct interviews (at baseline, 1, 4, and 7 months) and FGDs with participants in the cohort of AGYW and ABYM (at 1, 4, and 7 months) and key informant interviews (KIIs) with stakeholders including district department of health officials, facility managers, HIV program managers and managers of non-governmental organizations working on the HIV programs (at 6 months).

### Data collection

Baseline quantitative data collection will be conducted using self-administered electronic questionnaires built into REDCap. During this exercise, we will collect demographic information including age, sex, employment status and level of education attained. We will also collect behavioral and HIV risk-related data, including the number of sexual partners, partner HIV status, condom use, recreational drug use, alcohol use and post-exposure prophylaxis. Table 4 gives an overview of variables, process indicators and data sources and tools, and how these relate to the objectives for this study. Questionnaires will be administered at 1, 4- and 7-month follow-up visits to assess factors influencing engagement with oral PrEP (Fig 2).

**Table 4:** Roles of outcome variables in the SHeS’Cap-PrEP study in uMgungundlovu district, KZN, 2021.

| Domain | Examples of variables | Examples of process indicators | Data sources and tools |
| --- | --- | --- | --- |
| Proportion of adolescents testing HIV-ve who successfully initiated PrEP immediately after HIV-ve diagnosis (post baseline)<br>(primary outcome) | Patient receipt of HIV test result.<br>Patient attendance at first clinic appointment after receipt of HIV result. | Proportion of newly diagnosed HIV-ve adolescents who initiated PrEP on day of diagnosis.<br>Time in days to receipt of HIV-ve result.<br>*Mechanisms of referral | Routine data (Clinical charts, TIER.Net*) |
| • Proportion of adolescents continuing in PrEP 1-, 4- and 7- months after negative HIV diagnosis<br>• Proportion of adolescents who indicated for PrEP but did not continue in PrEP care,<br>• Proportion of adolescents who no longer indicated for PrEP, and<br>• Proportion lost to follow-up for PrEP care<br>• (primary outcome) | Patient attendance at SDPs as per PrEP guidelines between 1-7 months of a negative HIV diagnosis. Patient collection of drugs as per guidelines. | Utilisation of health information systems including patient tracking and follow-up processes, defaulter lists generated (through TIER.Net or other methods) | Routine data (Clinical charts, TIER.Net) |
| Social & demographic variables associated with PrEP initiation and continuation at 1-, 4- and 7-months | Age, gender, marital status, ethnicity, level of education, level of poverty | Quality and completeness of facility and TIER.Net records when triangulated against questionnaire data | Routine data (Clinical charts, TIER.Net), Baseline and follow-up questionnaires |
| Further description of participants' health seeking behaviour | Participant experiences of seeking HIV testing and prevention prior to date of testing in this cohort. Proportion of patients who discontinued PrEP after initiation | Quality and completeness of facility-and district-level routine data. Extent to which facility-level records can be linked to district-level TIER.Net | IDI and FGD guides |
| Clinic processes and data quality to support PrEP initiation and continuation.<br>Association between SDPs processes and PrEP initiation and adherence | Completeness and location of post-test initiation (clinic/community/schools)<br>Proportion initiating PrEP immediately after a negative HIV test at SDPs | Provision of a letter/other written communication to facilitate referral at each of the SDPs. Proportion given appointment date; proportion referred to support group. Usage of paper-based and electronic registers and records systems (TIER.Net) at different SDPs to check results before/during patient appointments. Availability of PrEP, waiting times etc. | Routine data (Clinical charts, TIER.Net), Baseline and follow-up questionnaires |

*\*TIER.Net is an electronic patient management system used to capture patient-level data on HIV management in South Africa*

**Figure 2:**
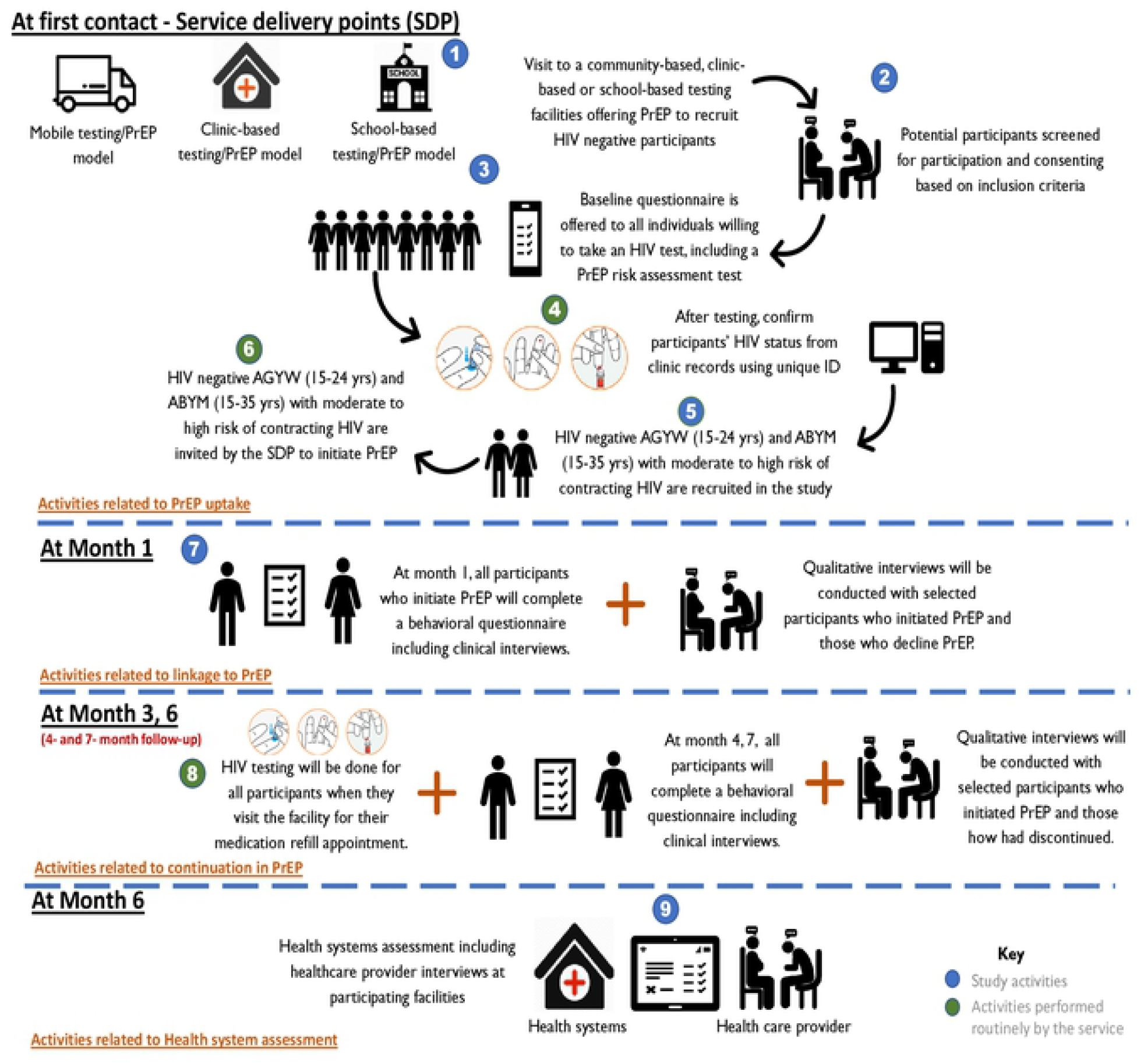
An illustration of the study activities for the SHeS’Cap-PrEP study in uMgungundlovu district, KZN, 2021

We will also use secondary data sources, including the district health information system data to obtain information on PrEP use and (dis)continuation in the SDPs where this project will be implemented. Information on the processes and procedures of the PrEP-based HIV prevention differentiated models will also be obtained from standard operating guidelines and other grey literature.

We will also collect qualitative data using IDIs, FGDs and KIIs. FGDs and IDIs will explore participants’ perceptions regarding the use of oral PrEP, including the barriers and facilitators for oral PrEP uptake, continuation, and adherence during oral PrEP use. We will conduct two FGDs (8-10 participants in each group) from each of the 4 groups (i.-initiated PrEP within 1 month), ii. Did not initiate PrEP within 1 month, iii. continued PrEP at 4/7 months and iv. did not continue PrEP at 4/7 months). We will conduct 12 IDIs from each of the four groups to explore information on how participants incorporated daily pill-taking into their daily routines, as well as obtaining information on security, privacy, and disclosure of PrEP use to partner(s), family, and/or friends. IDIs and FGDs will also be used to explore experiences of taking PrEP, including side effects and how it influenced participants’ decisions to either continue or stop. These IDIs and FGDs will also explore perceptions of, and experiences with the different models through which participants are receiving their PrEP. This will include exploring information related to participants’ experiences when using the model and how useful the model was to enhance their engagement in PrEP.

Furthermore, 18 KIIs will be conducted with all or purposively selected stakeholders including reproductive and health care providers at different sites and primary health care centers. The KIIs will focus on the barriers to, and facilitators of PrEP delivery, ease of integration of PrEP with other reproductive and health services, and perspectives of ways to improve the PrEP delivery models. During the KIIs, process mapping will also be conducted to understand the processes and steps involved in the delivery of the services in each of the models. In this case, emphasis will be placed on the inputs, outputs, and outcomes. The duration of the IDIs, FGDs, and KIIs will range from 60-120 minutes depending on how the discursive and interactive process unfolds.

### Data management

Fieldwork supervisors will do daily quality checks on completeness of REDCap records on the tablets, before uploading data over 3G or Wi-Fi to the REDCap folder stored securely on the SAMRC’s server. The project coordinator will also undertake daily checks for completeness and quality of REDCap records uploaded by each participating facility’s data champions, before confirming to the data champions at each facility which REDCap records can be erased from the facility-based tablets. All data will be quality controlled by the project coordinator. Inconsistencies and errors will be flagged at regular quality control meetings for data champions, and will remain on a watch list until corrected, with additional training supplied, if necessary, monitored by the project coordinator under oversight of the project manager.

#### Routine data from district-level TIER.Net

The facility-based TIER.Net database, an electronic patient management system used to capture patient-level data on HIV management in South Africa, will be used. After assessing readiness during the pre-implementation period, if district health staff are willing to engage in this, study staff will triangulate TIER.Net dispatches from participating facilities with district-level TIER.Net, seeking to identify any enrolled participants lost from care in their original facility who may be accessing care elsewhere in the district. This will be undertaken in a private area within the district health office and no TIER.Net patient-identifiable data will leave the district health office.

#### Qualitative data

Interviews will be audio-recorded and transcribed verbatim in the language in which the interview was conducted, then translated into English by accredited translation companies. To maintain quality control the transcripts will be assessed for correctness and accuracy in translation by bilingual project managers and fed into Atlas.ti version 10 (33) for analysis. A feedback workshop will be held with those who conducted the interview and those who participated in the interviews to ensure the transcribed information were correctly captured. All qualitative data will be coded manually, before being analysed more formally within Atlas.ti software. Interviews with health providers will be conducted in English.

REDCap data will be uploaded daily (identified only with enrolment number), using 3G or Wi-Fi as available, from facility-based tablets to the SAMRC-hosted REDCap server, which is password protected. All folders on the SAMRC file servers are additionally password-protected, as they can be accessed only by SAMRC staff members’ logons. This project will be assigned a specific folder, for which individual permissions will be assigned to specified staff (SAMRC co-investigators in this core evaluation team) to access them. Qualitative data (health systems assessments, interview audio files and transcripts) will also be stored in this project’s restricted access folder on the SAMRC secure server.

All hard copies of data (e.g., transcripts) will be filed and securely stored in a locked cupboard in SAMRC offices (normally the PI’s, occasionally in the offices of co-investigators, although digital copies will be accessed where possible). Recordings of qualitative interviews will be uploaded and stored (digitally within Atlas.ti and in paper form) by enrolment number, without personally identifiable information, into the password protected project folder on the SAMRC server. These will be destroyed after 15 years to ensure confidentiality.

### Data analysis

At baseline, demographic and behavioral characteristics will be analyzed, stratified by PrEP initiation. Since most of the variables will be categorical, proportions and percentages will be reported with their respective 95% confidence intervals. Continuous data will be reported as means (±SD)/medians, interquartile range (IQR) depending on data normality. The association between the demographic/behavioral characteristics and the stratification by PrEP initiation (or not) will be assessed by Chi-square/Fishers exact tests depending on the large sample size assumption or t-test/Mann Whitney test depending on data type (categorical and numerical, respectively).

Further analysis on the sociodemographic and behavioral characteristics amongst those who take up PrEP will be done during the follow-up times (t_1_, t_3_, t_6_). Uptake of PrEP or subsequent continuation will be reported as the number of adherent participants as a proportion of the baseline study sample (HIV negative diagnosed participants) or those that take up PrEP respectively. Adherence/Uptake will be defined as taking four or more daily pills in the previous seven days, according to patient self-report at each clinical follow-up visit. The different service delivery models will be compared based on gender and age; however, it is unlikely there will be initiation and continuation of PrEP at facility-only or community-only sites, but a hybrid. Hence, a descriptive analysis will be conducted, and where possible a statistical association between adherence and different service delivery points will be conducted at the cluster level.

We will also do descriptive analysis of the patterns of oral PrEP use presented as the time to initiation, duration of use and behavioral characteristics including contraception use, pregnancy, sexual activity and HIV incidence. Furthermore, hazard proportional modelling of time to discontinuation of oral PrEP will be attempted using the collected variables considered to be enablers or barriers to uptake/adherence to PrEP (including experiences of violence). A p-value of <0.05 will be considered as statistically significant for all tests.

Using the Atlas.ti qualitative data management software, the team will code the transcripts to identify emerging themes to address the objectives of the different work packages. First, a codebook for each objective will be developed and the identified coders will code the data independently. Discursive meetings will then be held among the coders whereby codes will be modified, collapsed, expanded, or dropped as needed. The data analysis process will be documented including the discussions and decisions around data analysis.

### Ethics and dissemination

Ethical approval for this study was obtained from the South African Medical Research Council Health Research Ethics Committee (Ref #: EC051-11/2020) on 19 January 2021. Gatekeeper permissions were also obtained from the KwaZulu Natal Provincial Departments of Health (Ref #: KZ_202010_033), the uMgungundlovu health districts, and facilities. This project was reviewed by the U.S. Centers for Disease Control and Prevention (Atlanta, GA), Centers for Global Health Associate Director for Science in accordance with CDC human research protection procedures and was determined to be research, but CDC investigators did not interact with human subjects or have access to identifiable data or specimens for research purposes. A waiver of parental consent has also been granted for participants aged 15-17 years, and documentation and informed consent process will be followed as for participants 18 years and older.

We will obtain oral and written informed consent from all potential participants in the study prior to their participation. Participation will be voluntary, and participants will be informed during recruitment that they could withdraw at any stage without consequences from the study team or the facilities which they attend. Any event deemed to be a serious adverse event (e.g., breach of participant confidentiality) will be systematically recorded and will follow the Standard Operating Procedures of the SAMRC Ethics Committee, which includes reporting adverse or unexpected events within 48 hours to the SAMRC Ethics Committee. Besides the emotional impact of discussing personal matters, adverse events are unlikely, as this evaluation carries minimal risk (according to the CDC Consent Form Template and definitions).

Dissemination of results will be extensive, both through scientific fora (publications and conference presentations) and through feedback and materials for the department of health, at facility, district, and national levels. The findings of the study will be presented to facility-based, district, provincial and national stakeholders, implementing partners, researchers in the scientific community and members of the public of South Africa, including participants attending primary care facilities in uMgungundlovu district. The anonymity and confidentiality of the participants will be preserved by not revealing any identifying information during the dissemination of study findings. Outputs will include a brief policy-relevant summary of findings, peer-reviewed journal articles, research capacity building through one or more research degrees, and materials to support enhanced capacity for scale-up of the interventions.

## Discussion

This study aims to identify and enhance the empirical understanding of the effective and feasible PrEP models of care for improving PrEP uptake, continuation, and adherence among AGYW and ABYM in uMgungundlovu district, KwaZulu-Natal province, South Africa. The South African government have implemented differentiated care models to address continued high HIV incidence among AGYW and ABYM, and the factors that increase their vulnerability to infection. PrEP has several advantages over other HIV prevention methods for women at high risk of contracting HIV including autonomous or covert use and not requiring use at the time of a sexual encounter (34, 35). Many factors contribute to lack of adherence and continuous use of PrEP among AGYW, including lack of knowledge, partner support, intimate partner violence (IPV), the need to conceal use, limited private storage space, fears of inadvertent disclosure, subsequent misperceptions about HIV serostatus, routine disruptions and not having a dose available (36-38). Inadequate PrEP knowledge has been shown to be one of the main barriers to PrEP use among ABYM (39).

Findings from this study will provide policymakers and implementers with information on the rates of PrEP uptake, models of PrEP delivery that are most successful in improving PrEP uptake, continuation, and adherence especially for at-risk populations such as AGYW and ABYM in South Africa. It will also describe the HIV testing experiences and the social, economic, health system, and behavioural factors that facilitate or impede PrEP cascade outcomes, including factors relating to acceptability of service delivery models for AGYW and ABYM. These will allow stakeholders to implement focused interventions and target populations who require additional effort to implement PrEP based HIV-prevention strategies for these population.

The strengths of this study include the combination of research methods to answer the research questions. This study will provide critical process information and will identify best service delivery models for successful initiation and continuation in PrEP for the target population. It will also provide South African policymakers and implementers with information on the rates of PrEP uptake, which models of PrEP delivery and care are preferred and work, and for whom they work. This will allow stakeholders to implement focused interventions and target populations who require additional effort to implement PrEP based HIV-prevention strategies.

The possible limitations of this study include selection bias in that we are sampling adolescents and young people accessing the different service delivery points (facility, community, and schools), this bias will likely be present as adolescents with severe physical or mental health conditions, carer responsibilities or those from lower-socio-economic backgrounds, may be absent on the day of recruitment, particularly during the SARS CoV-2 pandemic. Due to COVID-19-related disruptions to the health system, clinic records may not be updated with the latest clinic or laboratory data for patients, with missing clinic visit history details for patients who missed appointments during this period. We will work with health care providers to flag missing patient data and work with them to assess how these data could be retrieved. Furthermore, due to the challenges with reaching participants in this setting for follow-up interviews, our window period to assess test-retest reliability is large (within two weeks of post-baseline completion). However, we will initiate follow-up interviews within 14 days post-baseline questionnaire completion to reach the bulk of participants within a suitable timeframe.

## Data Availability

No datasets were generated or analysed during the current study. All relevant data from this study will be made available upon study completion.

## Authors’ contributions

EN and CM conceptualized the study. MH and EN drafted the manuscript. EN, CM, WB, CL, JB, MC, SA, and SD provided methodological support, while EN, CM, TR, WB, MH, DG, TM-A, NJ, NF, DP, SA, AP-V, JD, MC, SD and JB provided critical contributions towards developing and refining the manuscript. All authors read and approved the final manuscript.

## Disclaimer

The findings and conclusions in this report are those of the authors and do not necessarily represent the official views of the Centers for Disease Control and Prevention.

